# Cerebrospinal Fluid Proteomics Reveals Inflammatory Activation in Aneurysmal Subarachnoid Hemorrhage Irrespective of HIV Status

**DOI:** 10.1101/2025.09.30.25337004

**Authors:** Rohen Harrichandparsad, Gil Lustig, Victoria Sviridchik, Zesuliwe Jule, Mallory Bernstein, Kajal Reedoy, Yashica Ganga, Afrah Khairallah, Farina Karim, Vinod B. Patel, Ahmed Iqbal Bhigjee, Duncan Royston, Khadija Khan, Alex Sigal

## Abstract

**Objectives:** People living with HIV (PLWH) are at increased risk of cerebrovascular abnormalities, including aneurysmal subarachnoid hemorrhage (SAH). However, it is unclear whether HIV-associated inflammation contributes significantly to the inflammatory response observed in the cerebrospinal fluid (CSF) during aneurysm rupture. Here, we used high-throughput Olink proteomics to compare inflammatory marker profiles in CSF between participants with aneurysmal SAH and PLWH without aneurysms.

**Design:** This was a cross-sectional observational study which enrolled participants who were indicated for endovascular coil embolisation due to ruptured anterior communicating artery aneurysms (n=30) or undergoing clinically indicated lumbar puncture as part of workup for a non-neurovascular condition (n=9).

**Methods:** We performed lumbar puncture and analyzed CSF samples from individuals presenting with aneurysmal SAH (n=30) and PLWH without any known vascular pathology (n=9). Among aneurysm patients, 13 were PLWH and 17 were HIV-negative. An Olink Target 96 Inflammation panel was used to quantify inflammatory proteins.

**Results:** We assessed inflammatory profiles in cerebrospinal fluid (CSF) using Olink proteomics in individuals with ruptured anterior communicating artery aneurysms, with and without HIV infection. Among 68 detectable inflammatory proteins, 43 were significantly upregulated in participants with aneurysms (n=30) compared to people living with HIV (PLWH) without aneurysm (n=9). A similar inflammatory signature was observed in HIV-negative aneurysm participants (n=17) and PLWH with aneurysm (n=13), with no significant differences between these two groups. Interleukin-6 (IL-6) was the most upregulated protein across all aneurysm to non-aneurysm comparisons. These findings suggest that aneurysm rupture is associated with a strong CSF inflammatory response, largely independent of HIV status.

**Conclusion:** Ruptured intracranial aneurysm is associated with strong upregulation of inflammatory proteins in the CSF. This inflammatory response appears largely independent of HIV infection.

## Introduction

Cerebrovascular complications, such as aneurysmal subarachnoid haemorrhage (SAH), pose significant health risks, particularly for people living with HIV (PLWH), who are known to have an increased incidence of cerebrovascular abnormalities (1–3).

Aneurysms are thought to occur due to disruptions in wall shear stress (WSS) of blood vessels in the CNS, explaining their high tendency to occur at vessel branch points (4–6). Disruptions in WSS leads to endothelial dysfunction and a proinflammatory response mediated through NFκB (4, 5). These events have been shown to increase the levels proinflammatory mediators, including IL-6, a canonical activator of the NFκB pathway (7, 8) The extent to which HIV-associated inflammation influences the inflammatory response in CSF during intracranial aneurysm rupture remains poorly understood. Previous studies suggest that HIV infection may exacerbate systemic and central nervous system inflammation, potentially amplifying the inflammatory cascade triggered by SAH (9–12). To understand the effects of HIV status during intracranial aneurysm rupture, we recruited a cohort of participants indicated for endovascular coil embolisation due to ruptured anterior communicating artery aneurysms from Durban South Africa, an area with high HIV prevalence(13). We used Olink proteomics to characterize and compare inflammatory marker profiles in the CSF of individuals with ruptured anterior communicating artery aneurysms, with and without HIV infection, and PLWH without neurovascular pathology.

## Materials and Methods

### Informed consent and ethical statement

This was an observational study with longitudinal sample collection. The study protocol for blood and CSF and matched blood collection from participants indicated for SAH surgery was approved by the Biomedical Research Ethics Committee at the University of KwaZulu-Natal (reference BE480/15). Adult patients (>18 years old) presented at Inkosi Albert Luthuli Central Hospital in Durban, South Africa, Departments of Neurosurgery and Neurology. Written informed consent was obtained for all enrolled participants. CSF and matched blood were obtained from participants indicated for non-aneurysm associated lumbar puncture enrolled at Inkosi Albert Luthuli Central Hospital and King Edward VIII Hospital in Durban, South Africa after written informed consent (University of KwaZulu-Natal Institutional Review Board approval BE385/13).

### Participants

We analyzed CSF samples (1-5mL) and matched blood from individuals presenting with aneurysmal SAH (n=30) and PLWH without any known vascular pathology (n=9). Among aneurysm participants, 13 were PLWH and 17 were HIV-negative.

### HIV viral load

Viral load was quantified at an accredited diagnostic laboratory (Molecular Diagnostic Services, Durban, South Africa), using the RealTime HIV-1 viral load test on an Abbott machine.

### Olink proteomics

An Olink Target Inflammation panel was used to quantify 92 inflammatory proteins. Proteomic data obtained from the Olink platform were processed using the limma package in R. FDR correction was done using the Benjamini-Hochberg (BH) method. Proteins detected in fewer than 75% of samples were excluded, yielding 68 analyzable markers. In these proteins, any individual measurements below limit of detection (LOD) were set to the LOD value. Graphing of results was performed with ggplot in R.

## Results

We enrolled 30 participants with SAH, with 13 being PLWH and 17 HIV-negative (Table 1). Written informed consent was obtained for all enrolled participants. Enrollment was prior to the indicated surgical procedure and a CSF sample was taken at enrollment. In addition, 9 PLWH were enrolled clinical indications for lumbar puncture and characterized in a previous study (14). Median age was 42 the aneurysm group and 41 for the no aneurysm group, with no significant difference between them. Likewise, the proportion of males was about 40% and not different for any of the groups. The median HIV viral load was undetectable (threshold of detection = 40 HIV RNA copies/mL) for PLWH with and without aneurysm. However, there was a borderline significant difference in this parameter because there were 5 HIV viremic individuals in the aneurysm group (Table 1).

**Table 1:** Participant characteristics.

| Aneurysm |  |  |  |  |  |
| --- | --- | --- | --- | --- | --- |
|  | All | PLWH | HIV Negative | No aneurysm | p-value |
| **n** | 30 | 13 | 17 | 9 |  |
| **Age (Median, IQR)** | 42 (38,54) | 40 (36-54) | 42 (40,53) | 41 (40-50) | NS* |
| **Plasma VL (IQR)** | - | 40 (40-2469) | - | 40 (40-40) | 0.05 |
| **HIV viremic** |  | 5 |  | 0 |  |
| **Sex (male)** | 12 (40%) | 5 (39%) | 7 (41%) | 4 (44%) | NS** |

We quantified 92 inflammatory proteins in cerebrospinal fluid (CSF) from individuals with SAH and people living with HIV (PLWH) without vascular disease using the Olink Target Inflammation panel. After exclusion of proteins detected in <75% of samples, 68 markers were analyzable. Compared to PLWH without aneurysm, 43 proteins were significantly upregulated in the combined aneurysm group (Fig. 1a). A similar pattern was observed in subgroup analyses: 34 proteins were elevated in aneurysm participants living with HIV relative to PLWH without aneurysms (Fig. 1b) and 43 in HIV negative participants with aneurysm (Fig. 1c) relative to PLWH without aneurysm. Direct comparison between PLWH with aneurysm and HIV-negative aneurysm patients revealed no differentially expressed proteins (Fig. 1d).

**Figure 1:**
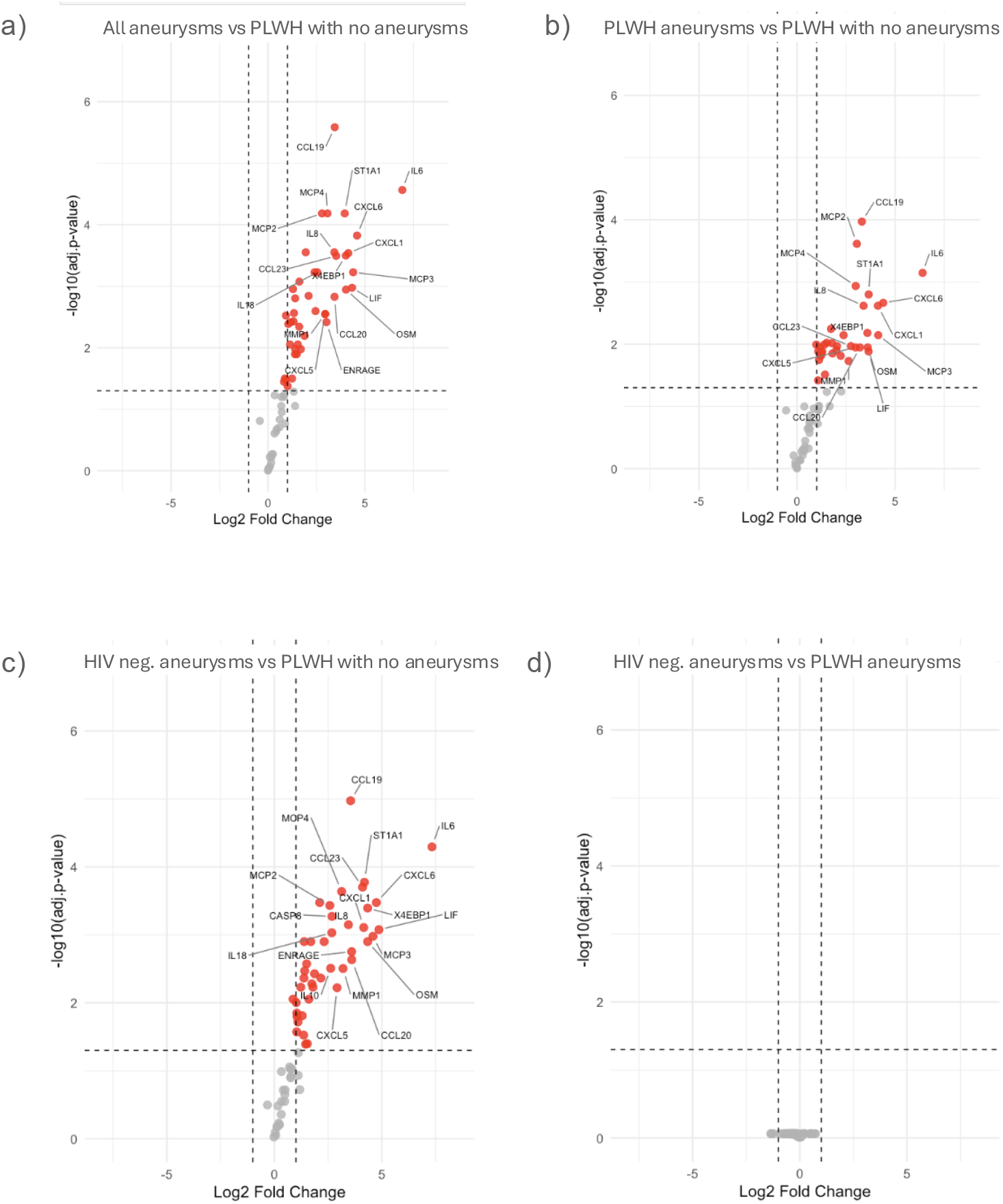
Volcano plots showing differentially expressed inflammatory proteins between groups. a) All aneurysm participants versus PLWH without aneurysm. b) PLWH with aneurysm participants versus PLWH without aneurysm. c) HIV negative aneurysm participants versus PLWH without aneurysm. d) HIV negative participants with aneurysm versus PLWH with aneurysm.

The top five most upregulated proteins in each aneurysm comparison showed marked fold-changes and were largely overlapping between subgroups (Fig. 2). NPX distributions for all significant proteins demonstrated consistent elevation across aneurysm patients, with top 5 upregulated proteins in the full aneurysm group versus non-aneurysm PLWH participants by NPX value being MCP1, CCL19, IL8, IL6 and CXCL1 by NPX value (Fig. 2a) and IL6, CXCL6, MCP3, LIF and CXCL1 by fold change (Fig. 2b). For PLWH with aneurysms versus PLWH non-aneurysm participants, top 5 upregulated proteins by NPX were MCP1, IL8, IL6, CXCL1, CCL19 (Fig. 2c), and by fold change were IL6, CXCL6, MCP3, CXCL1, and ST1A1 (Fig. 2d). For HIV negative participants with aneurysms versus PLWH non-aneurysm participants, top 5 upregulated proteins by NPX were MCP1, CCL19, IL8, IL6, MCP4 (Fig. 2e), and by fold change were IL6, LIF, CXCL6, MCP3, and OSM (Fig. 2f).

**Figure 2:**
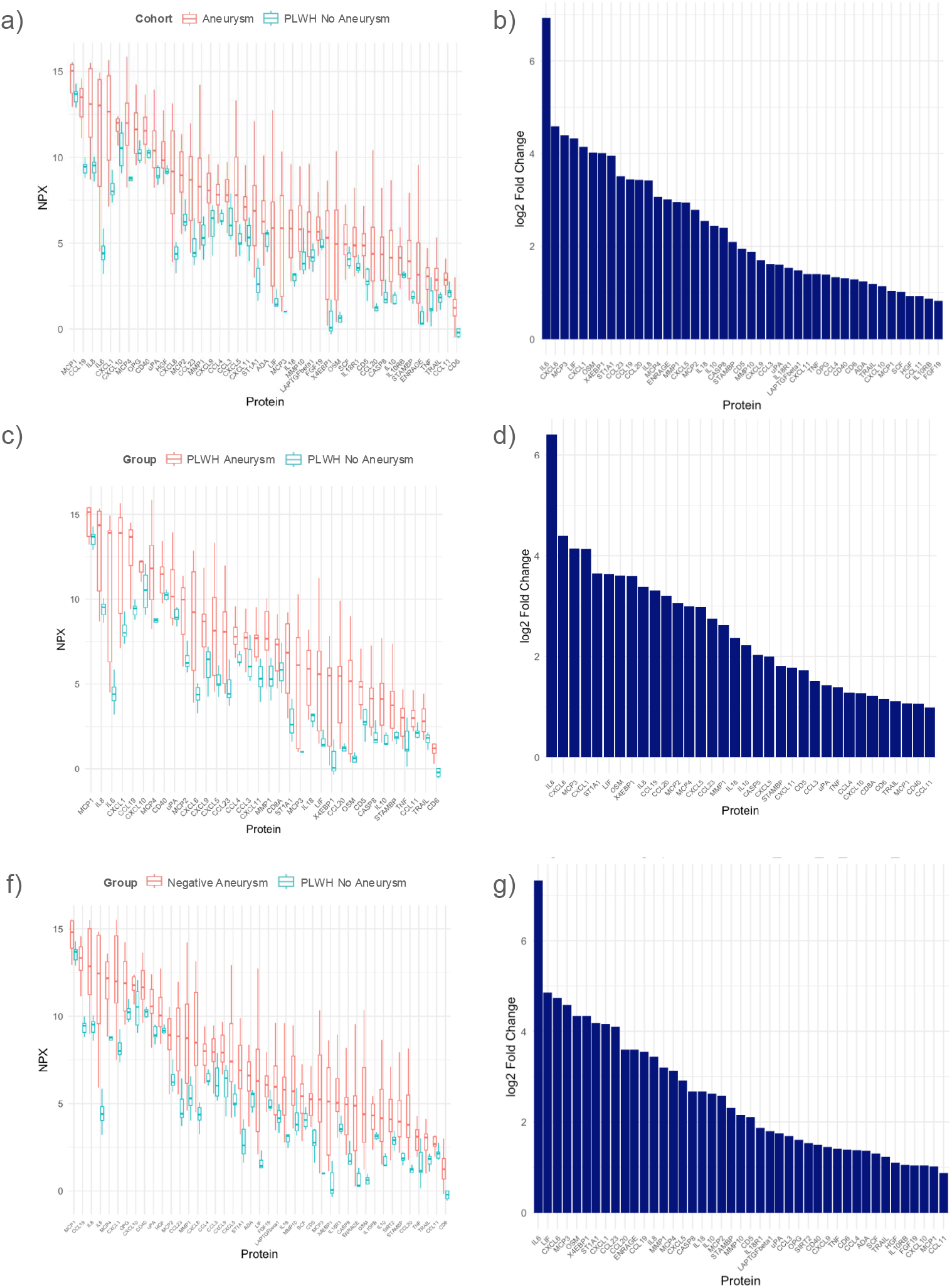
Absolute protein expression levels and fold changes for top markers in aneurysm vs. non-aneurysm comparisons. Log2 transformed normalized protein expression (NPX) and fold-change for: a-b) All aneurysm participants versus PLWH without aneurysm. c-d) PLWH with aneurysm participants versus PLWH without aneurysm. e-f) HIV negative aneurysm participants versus PLWH without aneurysm.

In all aneurysm-related comparisons, IL-6 showed the highest fold-change, followed by chemokines and cytokines including CXCL6, MCP-3, LIF, and CXCL1 (Fig. 2B, 2D, 2F). These findings indicate that aneurysmal SAH triggers a robust and consistent inflammatory response in CSF that is not substantially modified by concurrent HIV infection.

## Discussion

This study reveals a robust inflammatory signature in the CSF of individuals with aneurysmal SAH, characterized by upregulation of classical proinflammatory cytokines, most notably IL-6. Despite the known immune activation associated with HIV, PLWH with aneurysm exhibited a near-identical CSF inflammatory profile to HIV-negative aneurysm patients. This suggests that the local response to aneurysm rupture overrides any HIV-specific effects in the CNS compartment.

Limitations of the study include a small sample size and no HIV negative non-aneurysm group. Furthermore, there were several viremic participants in the PLWH aneurysm group while all PLWH without aneurysm were HIV suppressed in the plasma. However, given that the differences persisted between the HIV negative aneurysm group and the PLWH non-aneurysm group, it is unlikely that this HIV viremia in the PLWH aneurysm group is responsible for the pronounced difference in inflammatory markers.

Our findings demonstrate that aneurysmal subarachnoid hemorrhage elicits a robust pro-inflammatory response within the central nervous system, characterized by widespread upregulation of inflammatory mediators in the cerebrospinal fluid. IL-6 emerged as the most prominently elevated cytokine across all aneurysm-related comparisons, consistent with its established role as a key driver of neuroinflammation, glial activation, and secondary brain injury following CNS insults (15).

Our findings underscore the importance of inflammation in the pathogenesis and clinical course of aneurysmal SAH and suggest that HIV status does not substantially alter this response.

## Data Availability

All data produced in the present study are available upon reasonable request to the author

## Acknowledgments

We would like to acknowledge the CNS and SAH study participants and study staff.

## Author contributions

Rohen Harrichandparsad: Conceptualization /study design, data interpretation, wrote first draft.

Gil Lustig: Conceptualization /study design, data interpretation.

Victoria Sviridchik: Analyzed experimental data generated.

Zesuliwe Jule: Performed experiments.

Mallory Bernstein: Setup and managed the cohort and cohort data.

Kajal Reedoy: Performed experiments.

Yashica Ganga: Performed experiments.

Afrah Khairallah: Analyzed experimental data generated.

Farina Karim: Setup and managed the cohort and cohort data.

Vinod B. Patel: Study infrastructure, data collection, critical review and edits.

Ahmed I. Bhigjee: Conceptualization /study design, critical review and edits.

Duncan D. Royston: Conceptualization /study design, critical review and edits.

Khadija Khan: Conceptualization /study design, data interpretation

Alex Sigal: Conceptualization /study design, data interpretation, critical review and edits

## References

1. Thawani JP, Nayak NR, Pisapia JM, Petrov D, Pukenas BA, Hurst RW, et al. Aneurysmal vasculopathy in human-acquired immunodeficiency virus-infected adults: Imaging case series and review of the literature. Interventional Neuroradiology. 2015;21(4):441–50.

2. White EI, Anand P, Cervantes-Arslanian AM. Characteristics and evolution of cerebral aneurysms among adults living with HIV: A retrospective, longitudinal case series. Journal of Stroke and Cerebrovascular Diseases. 2023;32(6):107127.

3. Bulsara KR, Raja A, Owen J. HIV and cerebral aneurysms. Neurosurgical Review. 2005;28(2):92–5.

4. Chalouhi N, Ali MS, Jabbour PM, Tjoumakaris SI, Gonzalez LF, Rosenwasser RH, et al. Biology of intracranial aneurysms: role of inflammation. J Cereb Blood Flow Metab. 2012;32(9):1659–76.

5. Nixon AM, Gunel M, Sumpio BE. The critical role of hemodynamics in the development of cerebral vascular disease. J Neurosurg. 2010;112(6):1240–53.

6. Metaxa E, Tremmel M, Xiang J, Kolega J, Mandelbaum M, Siddiqui A, et al. High Wall Shear Stress and Positive Wall Shear Stress Gradient Trigger the Initiation of Intracranial Aneurysms. ASME 2009 Summer Bioengineering Conference, Parts A and B 2009. p. 523–4.

7. Braun DJ, Hatton KW, Fraser JF, Flight RM, Moseley HNB, Bailey CS, et al. Early changes in inflammation-related proteins in the cerebrospinal fluid and plasma of patients with aneurysmal subarachnoid hemorrhage. J Stroke Cerebrovasc Dis. 2025;34(6):108304.

8. Chen S, Hu Z, Zhao M, Sun J, Nie S, Gao X, et al. A comprehensive proteomic analysis reveals novel inflammatory biomarkers in intracranial aneurysms. J Proteomics. 2025;313:105374.

9. Valcour V, Chalermchai T, Sailasuta N, Marovich M, Lerdlum S, Suttichom D, et al. Central Nervous System Viral Invasion and Inflammation During Acute HIV Infection. The Journal of Infectious Diseases. 2012;206(2):275–82.

10. Sercombe R, Tran Dinh YR, Gomis P. Cerebrovascular Inflammation Following Subarachnoid Hemorrhage. The Japanese Journal of Pharmacology. 2002;88(3):227–49.

11. Chai C-Z, Ho U-C, Kuo L-T. Systemic Inflammation after Aneurysmal Subarachnoid Hemorrhage. International Journal of Molecular Sciences [Internet]. 2023; 24(13).

12. Naredi S, Lambert G, Friberg P, Zäll S, Edén E, Rydenhag B, et al. Sympathetic activation and inflammatory response in patients with subarachnoid haemorrhage. Intensive Care Medicine. 2006;32(12):1955–61.

13. Karim F, Gazy I, Cele S, Zungu Y, Krause R, Bernstein M, et al. HIV status alters disease severity and immune cell responses in Beta variant SARS-CoV-2 infection wave. Elife. 2021;10.

14. Lustig G, Cele S, Karim F, Derache A, Ngoepe A, Khan K, et al. T cell derived HIV-1 is present in the CSF in the face of suppressive antiretroviral therapy. PLOS Pathogens. 2021;17(9):e1009871.

15. Lucas SM, Rothwell NJ, Gibson RM. The role of inflammation in CNS injury and disease. British journal of pharmacology. 2006;147(S1):S232–S40.

